# An insight into birthing and EmONC service utilisation for making birthing safe: a qualitative study in underdeveloped districts of Punjab, Pakistan

**DOI:** 10.64898/2026.09.29.26364296

**Authors:** Shaheer Ellahi Khan, Muhammad Amir Khan, Nida Khan, John Walley

**Affiliations:** Health Services Academy, Islamabad, Pakistan; Association for Social Development, Pakistan; Nuffield Centre for International Health and Development, Leeds Institute of Health Sciences, University of Leeds, UK

**Keywords:** Skilled birth attendance, social determinants, lady health workers, birth planning

## Abstract

**Background:** The qualitative study was conducted to understand the socio-cultural factors (*challenges and enabling factors*) those influence the decisions and ability of the pregnant women and their families to make better use of the available safe birthing and EmONC services in two districts (Khanewal and Jhang) of Punjab, Pakistan.

**Objectives:** To explore the key social, personal and professional considerations (factors) that influence the women (and family) decision and ability to utilise various birthing and EMONC services and to understand care providers’ interest and ability to offer these safe birthing services in the districts of Jhang and Khanewal in Pakistan.

**Study Design:** The qualitative study was conducted, during the year 2013/14, on three rural health centers and two district headquarter hospitals in Kanewal and Jhang districts in Punjab. Through purposive sampling, 42 registered clients were interviewed to explain the different birthing experience along with 15 service providers, 2 district level programme managers and 2 transport vendors. The collected qualitative data from interviews were transcribed, and key emerging concepts/ themes were identified. A range and continuum of coded responses/ comments were compiled and discussed under each theme.

**Results:** Main themes were structured birth planning; antenatal at health centres; skilled birth attendance; emergency obstetric and neonatal care (EmONC); and EmONC travel. The study found that pregnant women consult their husbands and mother in laws for taking birthing decisions; consult the skilled birth attendant for management of complication; private clinics are preferred for various social and logistic reasons; many private clinics doing caesarean are not adequately equipped; male reluctance (for cultural reasons) to make in-advance transport arrangement for possible EmONC; travel voucher utilisation remained low because of limited coverage and less-friendly procedures.

**Conclusion:** Safe birthing decisions could be complex; and influenced by multiple social, cultural, economic, service access and quality considerations. Safe birthing practices can be improved if interventions remain sensitive to these preferences and realities.

## Introduction

International reassessment of program strategies to reduce maternal mortality show that most pregnant women are at risk of obstetric complications^a^. The high ratio of maternal mortality in Pakistan i.e. 276 deaths per 100,000 live births, indicates for a need to concentrate on the women’s health^b^. Social, cultural and economic factors play a crucial role in maternal deaths along with medical causes. The strategy adopted by most of the countries that have succeeded in overcoming the challenge of maternal and neonatal mortality, was to improve the access and utilization of skilled birth attendance and emergency obstetric care^2^. Pakistan has been striving to make the skilled birth attendance more available widely for pregnant women in relatively less-advantaged populations.

In Pakistan, the province of Punjab has an estimated population of more than 90 million.^f^ After devolution the responsibility of planning, financing as well as delivering health care lies with the provincial government. At district level the public infrastructure to provide health care (including MNCH), mainly comprises of: district and tehsil level hospitals; rural health centres; and basic health units. The main qualified skilled birth attendants in these facilities are doctors and lady health visitors (LHVs). In addition, the community midwives (CMW) are recently introduced community based workers whose key focus is to deliver safe birthing services. Lady health workers are general health promotion worker for bridging the links between clients and health care facilities. In each district, private health facilities are also providing birthing services which vary widely in cost and quality of care.

This qualitative study was conducted as a part of an intervention trial on promoting the use of safe birthing and emergency obstetric care services. The intervention trial led to an increase in skilled birth attendance in rural districts of Punjab. The trial showed an overall increase in skilled birth attendance from 64% (in control) to 75% (in intervention), without showing any reduction in neonatal and maternal mortality indices. National Bioethics Committee (NBC) Pakistan granted ethical approval for the intervention trial and its associated qualitative study (Ref No.4-87/10NBC-39/RDC/487). The trial was also registered with the Current Controlled Trials ISRCTN86264432.

This qualitative study was designed to explore the key social and personal considerations that influence the women (and family) decision and ability to use various obstetric and neonatal care options; and understand how do women cope with these social requirements. The study also focused on the key social and professional considerations (factors) that influence the care providers’ interest and ability to offer antenatal, normal delivery and caesarean section services.

### Rationale

Increasing numbers of randomized controlled trials of health interventions also include qualitative component^c^. The qualitative study was to help: a) assessing the socio-cultural and administrative feasibility of the intervention protocols; b) explaining the trial results/ experiences; and c) informing the intervention refinement for potential scaling.

### Methodology

A qualitative study design, based on in-depth interviews and focus group discussions, was used to study the client and provider experiences. The scope of the study included the: importance of counselling process, results of pre-birth interaction with skilled birth attendant(s), in-time preparation for delivery and EmONC (if needed), access to health facility, and socio-cultural perspective(s) in decision making process.

### Study area

The research was conducted in the two districts of Punjab i.e. Khanewal and Jhang, during period January – April 2014.

**Table 1:** Socio-demographic profile of two selected districts.

| **Indicator** | **Jhang** | **Khanewal** |
| --- | --- | --- |
| Estimated population (million) | 2.8 million | 2.5 million |
| Male (% of population) | 52% | 52% |
| Rural population | 70% | 80% |
| Total literacy | 37.1% | 45% |
| Women literacy | 21.4% | 52% |
| Population below poverty line | 24% | 32% |
| Average household size | 6.5 persons | 6.9 persons |
| Crude birth rate (per 1000) | 27.8 | 21.5 |
| Infant mortality rate (per 1000 livebirths) | 65 | 77 |
| Population growth rate | 2.16 | 2.4% |

### Sampling

The participants of the qualitative study included the registered clients of the trial, LHWs, skilled birth attendants, and EmONC staff. The sample was stratified into two main categories i.e., clients and providers. The mentioned categories were studied in three RHCs (rural health centres) and two DHQs (district headquarters) which were selected through purposive sampling method.

### Client’s sample

The sample of the clients was selected through purposive sampling method from three arm(s) of the trial. In each arm, the respondent were randomly selected from a list of 20 clients in each category, to represent the experience of skilled birth attendance by: a) doctors, b) LHV, c) CMW, and d) unskilled birth attendant; and EmONC services: e) used when indicated, f) not used when indicated, and g) used when not indicated.

**Table 2:**
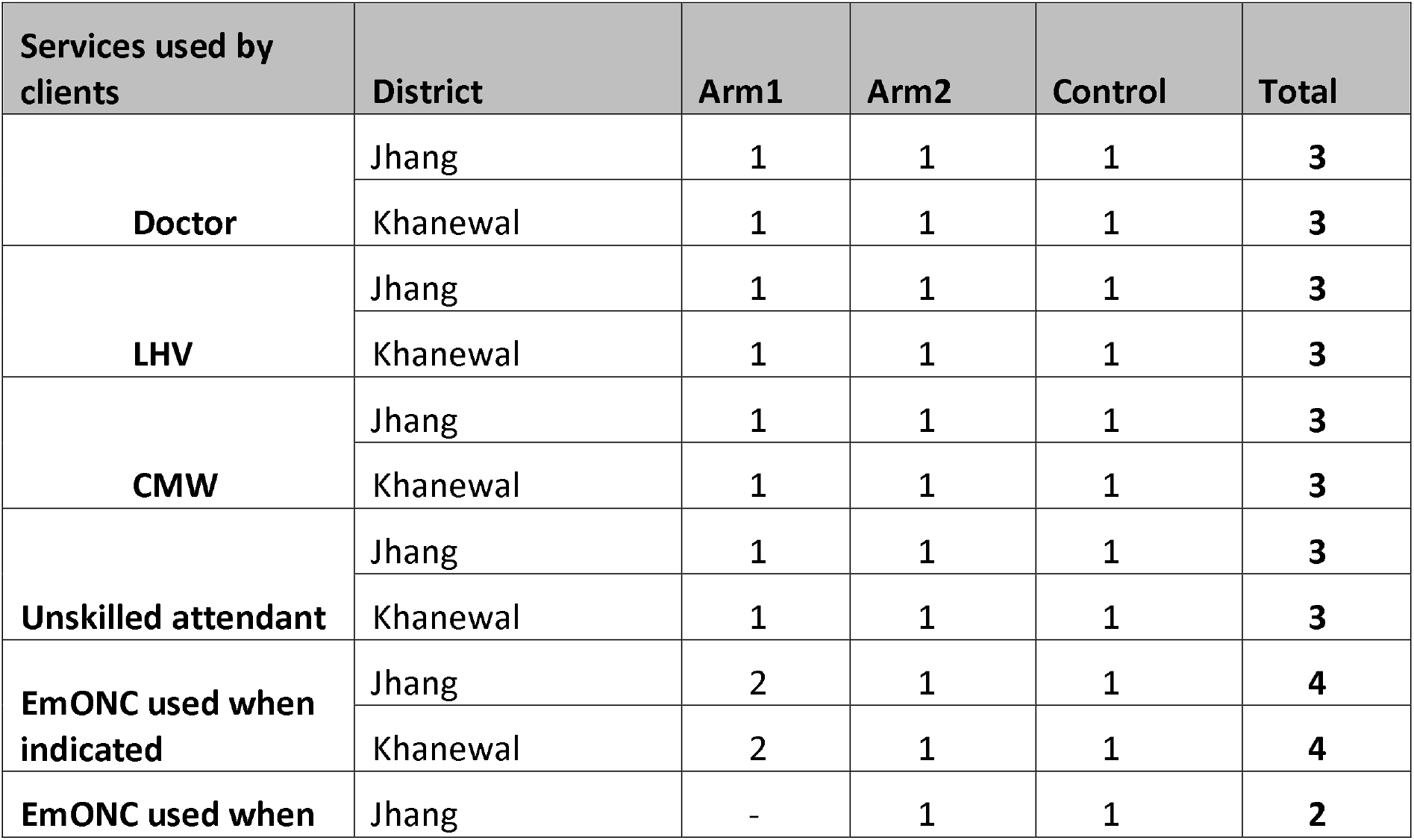
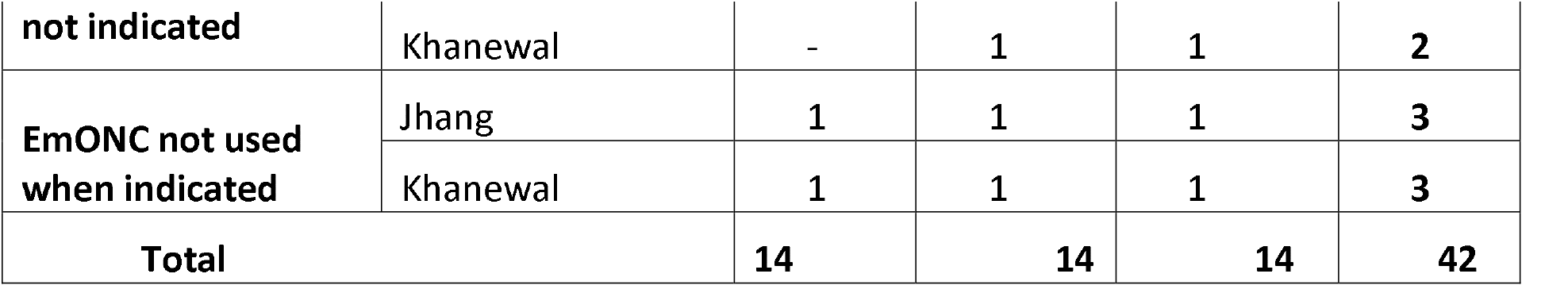
Details of the client sample.

A total of 42 interviews were conducted i.e. 14 each from two interventions and a control arms. In case of respondent non-availability, the next woman in the same list was contacted. A specially designed semi-structured interview schedule was used to interview women (and family) about their birthing experiences, including use of skilled birth attendance and EmONC services. The 38 selected respondents (women and families) were accessed, through respective lady health workers. The client responses were hand-recorded, with explanatory comments of the researcher. The collected data from interviews (both client responses and researcher comments) were transcribed, and key emerging concepts/ themes were identified. Then range and continuum of coded responses/ comments were compiled under each theme.

### Provider’s sample

The provider sample was purposively selected and was divided into three categories including:

- Lady health worker
- Skilled Birth Attendant/Doctor
- EmONC staff

**Table 3:** Details of the provider sample.

| **Provider** | **District** | **Arm1** | **Arm2** | **Control** | **Total** |
| --- | --- | --- | --- | --- | --- |
| **LHW** | Jhang | 1 | 1 | 1 | 3 |
|  | Khanewal | 1 | 1 | 1 | 3 |
| **SBA/Doctor** | Jhang | 1 | 1 | 1 | 3 |
|  | Khanewal | 1 | 1 | 1 | 3 |
| **EmONC Staff** | Jhang | 2 | 0 | 0 | 2 |
|  | Khanewal | 1 | 0 | 0 | 1 |
| **Total** |  | 7 | 4 | 4 | 15 |

A sample of 15 service providers was selected in two districts, through purposive sampling method, for semi-structured interviews. In addition two district level programme managers and two transport service providers were also interviewed in two districts. In each of the two district-headquarter hospitals, the staff engaged in delivering the comprehensive EmONC service was interviewed. A specially designed semi-structured interview schedule was used to interview the service providers about their respective experience of offering birthing services. The set of tools was piloted before using in the study.

### Data Collection

The data was collected by an expert team of field anthropologists. The selected respondents were contacted, through the respective lady health worker, for their consent and access. The interviews were conducted, either at the respective health facility or the respondent place, by a pair of trained qualitative data collection persons accompanied by the respective lady health worker. The client responses were hand-recorded, with explanatory comments of the researcher. A senior anthropologist supervised the data collection, by participating in few initial and then few random interviews.

### Data Analysis

The collected data from interviews (both client responses and researcher comments) were transcribed, using the rapid assessment technique. The key emerging concepts/ themes were identified. Then a range and continuum of coded responses/ comments were compiled under each theme. Both client and provider perspectives data were transcribed separately.

In consultation with the programme staff, the findings from various respondents were finally organized under the following major pragmatic areas of interest:

- *Structured birth planning:* covers mainly the client (and family) and the health services perspective of information-based decision making for safer birth planning including EmONC (if needed).
- *Antenatal Visits:* covers mainly the client and the provider’s perspective to antenatal visits for safe birthing.
- Skilled birth attendance: covers mainly the client preferences, circumstances, and experiences of skilled (or unskilled) birth attendance.
- *EmONC services and travel support: covers mainly* the client preferences, circumstances, and experiences of deciding and accessing the EmONC.

## Results

The findings of qualitative interviews with various respondents are outlined under five main headings: structured birth planning; antenatal at health centres; skilled birth attendance; EmONC services; and EmONC travel.

### Structured Birthing Planning

Most of the lady health workers found it manageable for them to gather and submit the skilled birth attendant and transport service provider data for the Directories. Mostly LHWs know about the available skilled (and unskilled) birth attendants and some transport providers in their respective populations. Most respondents suggested updating the Directories on every two-year basis.

LHWs reported to have conducted most of the birth planning session at pregnant women place, with only few at the health houses. Most LHW respondents reported that pregnant women do not/ cannot generally take the safe birthing related decisions without consulting their husbands and mother in laws for logistical-cum-monetary and social-cum-technical concerns respectively. This reliance on family member advice/ consensus is perceived to be more in rural women as compared to urban women. In general, the assumed ideal ‘man-wife’ decision for safe birthing is strongly influenced by mother-in-law and other senior members of the husband family. The study showed significant influence of mother-in-laws on birthing decision making. The reported reasons for women respecting the mother-in-law judgment on birthing matters include: relevant practical experience; holistic view of available resources and priority needs; trust of woman’s husband; in-depth cultural understanding; and valid negotiation experience. However, LHWs reported that some women do feel more able to openly discuss their practical constraints, when they are not accompanied by mother-in-law.

Furthermore, the LHW reported perceived reasons for an individual preferring the unskilled birth attendant (Dai) includes: elders’ historic experience; cultural preference; service package broader than just child delivery; perceived low-cost with flexible payment modality of Dais; and client lack of knowledge about availability and cost of skilled birth attendants.

Most respondent pregnant women found the LHW structured birth planning session useful. The most appreciated parts of the session were: a) LHW providing practical/ factual information in a usable form (e.g. provider contact, expected availability and cost etc.); and b) helping to make decisions and preparations for safe birthing. Most respondent women reported the inclusion of husband or mother-in-law in their birth planning exercise to be helpful. Most women felt that this sensitisation of family decision makers, through participation, helped in their selecting a skilled birth attendant rather than Dai (i.e. an unskilled birth attendant). Women reported the in-advance contact with selected skilled birth attendant to be useful, in terms of women feeling more confident about the availability and cost of the planned safe birthing services.

Respondent women with some history of obstetric complication (own or family member) were found more interested in planning for EmONC services, as compared to women with no history of obstetric complication. Many women respondents perceived the EmONC service to be a too technical matter for them to decide, without consulting their respective birth attendants. Most respondent women stated that they were encouraged to make some savings from the monthly family income, so that they can get the required EmONC services, if needed. Most respondent women also stated that their family did not arrange in-advance the transport for a possible travel to EmONC service.

The district level managers endorsed the importance/ need for an enhanced lady health worker ‘gate-keeping’ role to promote the utilization of available skilled birth attendants. This also postulates the requisite of follow-ups of all plenary and consultative sessions that may help in promoting safe child delivery services that will directly benefit the potential service users.

### Antenatal at Health facilities

It was found that pregnant women and their families were more enthusiastic about making antenatal visits if the woman (or a family member) had a history of complications in previous pregnancies. The respondent women revealed that the decisions whether? when? where? to visit for an ante-natal check-up need endorsement of mother-in-law. Few women also mentioned the challenge of finding time, out of their daily household responsibilities, to get an antenatal visit.

The respondent women revealed that an antenatal visit generally includes clinical examination and laboratory investigation (including scanning). Many of these services do cost money even if birth attendant at a public health centre in contacted. The reported cost items during antenatal exams at public facility were mainly: ultrasound scan (always done privately); and blood and urine tests. The respondents reported that the cost of antenatal is not very different for attending a skilled birth attendant at a public or a private facility.

“As we selected Baaji (LHV) for my delivery, so this made more sense also to get the antenatal care from her” (woman respondent: Jhang)

Many women respondents also mentioned other reasons for preferring care from a private clinic/ provider. These included: geographic proximity; relatively flexible clinic timing; service quality; and more supportive staff attitude as compared to public health centres.

“Baaji (LHV) who does not pay attention at a government centre, behaves differently when you visit her private clinic”. (woman respondent: Jhang)

The care providers at public health facilities, reported interruption in consumables required for antenatal exams (laboratory chemicals/supplies; glucose strips) and difficulty in maintaining the equipment (e.g. blood pressure apparatus; weighing scale). However, staffs were found reluctant to discuss issues around private practice and its relevance to the antenatal care being provided at public health centers.

### Skilled Birth Attendance

As mentioned earlier, respondent women had a general consensus about usefulness of information provided by LHW about availability, quality and price of the skilled birth attendants. However, some clients perceived that these recommendations are not always totally neutral.

“Sometimes LHW tries to promote a skilled birth attendant of her own preference, for reasons other than technical competence”. (Woman respondent - Khanewal)

Many of the LHW respondents also confirmed the women (and family) interest in getting actionable information about skilled birth attendants; and selecting the birth attendant that suits their circumstances (i.e. ability to pay; distance; cultural norms etc.). Many women also reported reliance on LHW recommendation for selecting among the available and feasible skilled birth attendants.

Many of the respondent women confirmed their visiting, usually with their mother-in-law, the selected birth attendant before the act of actual delivery. The mother-in-laws were reported to have discussed with the birth attendant their availability and place of delivery; also negotiated the service fees for the agreed delivery arrangement. In case of doctors, the client felt relatively less able to ask too many details or to negotiate delivery modalities, including payment. The respondent women also reported that they trust and comply, where possible, the recommendations of selected birth attendant.

“The doctor subsidised the service-fees, when she learned about our economic situation” (Woman respondent: Jhang)

Some respondent women also reported incidents of changing the initially selected skilled birth attendant because of: non-satisfaction with prospects of the service availability at odd times, if needed; not-agreement on home delivery; and non-agreement on service payments.

The respondent doctors and lady health visitors confirmed that staff appointed at public health facilities do private consultation in the evening; doctors practice family medicine whereas LHV practice in mother and child health care. These respondents also admitted that some of the privately practicing doctors and lady health visitors do take decisions that are difficult to justify on medical grounds. These respondents also confirmed that many women (and family), when come to the private clinic, do enquire about clinic (service) timing and ability to deal with possible complications, including availability of required equipment; also negotiate the service charges.

The respondent lady health visitors revealed that many of the women who come for their antenatal check-up end-up getting delivered by the same lady health visitor. They also mentioned their working linkages with lady health workers in the area, and LHW useful role to inform pregnant women about the importance of skilled birth attendance. They have revealed weak district arrangements to supervise their safe birthing practices.

Some women respondents remarked that doctors and lady health visitors are more easily available at their private clinics as compared to the respective public health facility. Some women also commented that private providers are more available for odd-time delivery (i.e. night) as compared to the public facilities.

The women respondents reported that community midwives are relatively more available for home-based deliveries as compared to lady health visitors; whereas doctors are only occasionally available for home-based delivery for a very special reason, situation or a person. Lady health visitors were also reported to provide home-based delivery but this generally costs higher, especially if conducted at odd-times. The women respondents also reported that generally skilled birth attendants provide the materials used in conducting delivery, and subsequently charge to the client.

Women respondents mentioned few social reasons for people preferring unskilled birth attendants. These include: historic presence in the community; low-cost with flexible payment modalities; home-based delivery; more comprehensive social service package including sharing household work during immediate pre and postnatal period (when women is less able to carry out such activities). One woman respondent also mentioned that because of mother-in-law strong views in favor of unskilled birth attendant, she was not able to opt/ use the skilled birth attendant.

LHW respondents mentioned unskilled birth attendant’s reaction to their (LHW) promoting the skilled birth attendants. Some of the Dais (unskilled attendants) even tried to counter the LHW (gate-keeping) efforts by misinforming the elderly women about skilled birth attendance and even challenged lady health worker intention/ character.

The district level managers interviewed endorsed the reality that many doctors and lady health visitors at public health centers are engaged in private practice. They agreed that public health centre doctors/ LHV do advise women additional investigations not currently available at respective centers. They also acknowledged district limited ability to maintain equipment at public health facilities; and also to influence the quality and price of care being offered at private clinics. The managers also indicated the need for further strengthening the refresher training and supervision of community-based midwives and facility based lady health visitors for improved quality of birthing practices.

### EmONC Services (Emergency Obstetric and Neonatal Care)

Many respondent women recognized the importance of their (and family) knowing the signs of danger in pregnancy. However, most women admitted that getting operated is too technical a decision for them to take, so mostly rely on an advice of the selected birth attendant for their decisions.

“When I faced vaginal bleeding (before birth); I called Baaji (LHV) to ask what should we do?” (Woman respondent Khanewal)

All women reported to have gone for an operation (caesarean) on advice of the respective care provider (i.e. doctor or lady health visitor). No respondent mentioned getting operated (caesarean) by choice. Women also mentioned that the first pregnancy makes people more sensitive/ apprehensive about getting the safe birthing care. Women generally agreed that getting a caesarean is a provider-driven decision, with very limited (if any) say of the client other than to agree/ comply.

Many women could state the possible complication they experienced that resulted in their caesarean. Two main complications these women mentioned were: vaginal bleed and prolonged labour. However, many women mentioned that they got operated, without experiencing a fit or vaginal bleeding or prolonged labour, because they were so advised by the respective care provider (i.e. doctor or lady health visitor).

“Care providers do sometimes exaggerate the risk of a possible complication to convince their client for a caesarean”. (LHW respondent - Jhang)

Some women who did not get a caesarean, even when indicated, held the respective birth attendant responsible for not advising them to get the caesarean (EmONC).

“I did get longer labour at her private clinic, but Baaji (LHV) kept assuring me that there was nothing to worry” (woman respondent - Jhang)

Both women and lady health workers mentioned that for many clients the district level public hospital is not a preferred source of EmONC services mainly because difficult geographic access; perceived lack of attention; perceived lack of care quality; and the cost not very different from the private clinic.

Many of the lady health workers and some women respondents agreed that many of the private clinics/ hospitals conducting caesarean seems not adequately equipped with qualified staff and equipment for the operation. These respondents also mentioned that many lady health visitors are engaged in conducting (unauthorized) caesarean at private clinics, by falsely claiming the presence of a qualified doctor for the caesarean.

Some of the doctors and lady health visitor respondents reported that some clients reach them at a relatively advanced stage of pregnancy complication, so they are left with limited option to refer such cases to an EmONC facility. Lady health visitors also mentioned availing the services of qualified doctors to deal with EmONC cases at their respective private clinics.

The district level managers endorsed the reality that few public hospitals are not enough for providing EmONC services in a district. However, they also agreed that there are no private clinics with basic requirements in-place (qualified staff; equipment) to offer comprehensive EmONC care. They also mentioned that some private clinics are engaged in offering EmONC services beyond their lawful capacity. Managers in districts suggested strengthening of the existing partnership mechanisms, so that private clinics can be brought into the mainstream provision of quality care.

### EmONC Travel

Both women respondents and lady health workers mentioned the reluctance of male members to talk in public or to outsiders about a private matter i.e. pregnancy in their household. The male members were reported to find it difficult to contact in-advance a transport; vendor; explain the possible (uncertain) need of EmONC travel; and negotiate a transaction that is not yet certain to happen. Furthermore, most families felt confident about their ability to arrange a transport locally, when needed.

Most women and lady health workers reported that EmONC transport arrangement, at a relatively short notice, is generally not a challenge; as most localities have multiple local vehicles and an individual transport owner (vendor or a neighbour) is culturally expected to respond positively to any such emergency need. The respondents did not report any incident where transport owner had refused or over-charged (exploited) the client emergency (i.e. EmONC need).

Interview with travel vendors revealed that they do feel morally and culturally obliged to help an individual who is in need of an emergency medical travel, more so if the person belongs to their own neighbourhood. Generally people know about the travel services operating in their respective neighbourhoods. The respondents revealed that in routine the travel cost is determined on the basis of travel distance, road condition, and travel timing (i.e. day or night).

“When we find someone in emergency (life is at risk) we do offer as much support (subsidy) as we can afford”. (Vendor Jhang)

The respondents also endorsed the cultural reluctance of men to discuss the private matter (pregnancy) openly; also relatively fewer men making an advance contact for a possible EmONC travel. The vendors also revealed that mobile phones have significantly improved their communication with clients; also claimed their reaching the client (in emergency) quite quickly as mostly in the same neighbourhood.

The reasons for low uptake of travel voucher, as reported by respondent women and lady health workers included long-time between the provision and actual use of the voucher resulted in many women loosing, damaging, or even forgetting the travel voucher (voucher was provided at the time of pregnancy registration); missed to carry the voucher along when they went for emergency services; timing of EmONC voucher reimbursement did not match their schedule; support documentation requirement was found not easy to manage; and voucher was not valid for the private facilities/ hospitals used by many clients for EmONC services (only valid for care at programme-endorsed facilities).

## Discussion

The pregnant women dependence on her family members for selecting the provider of birthing services is similar to many other developing country experiences. The location and plan of birth was found to be decided mainly by “men” and “elder women” in some sub-saharan^3,4,5,6,7,8^ and south asian^9^ countries respectively.

The rural domestic setup inclines one’s family to opt for traditional birth attendant keeping in view prior experiences, family and social ties, and avoiding gossip. On the other hand clients were interested to know and discuss about the skilled birth attendants in isolation with LHWs, as traditional values in many countries of South Asia prohibit discussion about reproductive health matters openly or between the sexes. Even if a woman or any of her family members were aware that one of the danger signs of a pregnancy was being experienced, the open acknowledgement of the emergency or the discussion on which course of action to be taken may be put off as long as possible, or not occur at all. ^10^

The present study showed similar indicators of negotiation trends with skilled birth attendants with a baseline in Bangladesh. Mothers who did not have skilled care attendants or did not use health facilities (public or private) for delivery care were asked to give reasons for not doing so and the commonly cited reason was the cost considerations, because they thought the cost for such care was high and unaffordable^11^. Whereas, having a skilled birth attendant (SBA) at every delivery has been found to markedly reduce maternal morbidity and mortality in countries such as Malaysia and Sri Lanka^12,13^. Studies have indicated that women who are birth prepared are more likely to be assisted by skilled birth attendants^14,15,16^. Observations have been made that those countries, which have managed to reduce maternal mortality drastically in the last 50 years, such as Malaysia and Sri Lanka, have invested much in increasing the number of SBAs and access to emergency obstetric care^17^. On the other hand traditional birth attendants attended the clients at home, and also provided additional services such as cleaning of the house and washing of clothes. The service package offered by TBAs could explain partly the reasonable uptake of unskilled birth attendants. Exacerbating the situation are low literacy rates, limited exposure to the world outside, and heavy domestic responsibilities which prevent women from proactively seeking health information themselves. Many are not even aware of what services are available.^18^

Clients visited the private health facility for antenatal visits while arguing about the less supportive and time taking mechanism at public health facilities including shortage of necessary supply. In the private health sector such services are expensive, while government health services for antenatal care are available at very subsidized rates. In another study sample, a very small proportion (14.5%) of women received antenatal care from government facilities, while a much higher proportion of women received care from private facilities ^19^, which is consistent with the findings of this study.

On the other hand most of the clients were unable to decide for antenatal visits without their respective mother-in-laws. According to a recent survey of women between the ages of 15 and 40 years in rural Punjab, only 28% can go unescorted to the local health center and only 12 percent can travel alone to the nearest village. Over two-thirds of the women interviewed required an escort to leave home.^20^

Mother-in-law and peer groups are helpful in developing traditional knowledge of learning about the danger signs/symptoms. The transportation was arranged when required and motorcycle Rickshaw was mostly used in the time of need, if required a small car was also arranged and in such times community supported the client and her family. Low awareness of the required steps towards a safe delivery can lead to unhealthy behaviours. Not making prior transport arrangements and therefore having to use uncomfortable transport like a rickshaw or *quingqi* are reflective of poor preparedness and have been reported by others.^21^

The financial challenges faced by families play vital role in taking decisions about the transportation arrangements. It was discovered that only a few clients used the travel voucher when required. On the other hand, findings from the maternal health voucher scheme in Bangladesh reveal that there was a significant increase in the utilization of delivery care at the facility but it was also observed that use of public-sector facilities for delivery services increased in only intervention sites while control sites experienced greater increase in using private sector.^22^

C-Section was mostly provider driven and clients were normally told in the sixth month of their pregnancy regarding the C-Section. Studies reflect that patients may leave the health-care related decision to the physician, because they trust his knowledge and expertise.^23^

The doctors at private health facilities did propose about the C-Section at least two to three months earlier. A qualitative study that was conducted in Lebanon in 2007 showed that lack of information about the post-partum period following the C/S operations affect women decisions of the route of delivery.^24^ Less educated client was an easy case to convince for a C-Section especially the clients with first pregnancy were more sensitive as compare to the clients who had experienced C-Section in previous pregnancies. A qualitative study showed that the most important reason for requesting C/S operations was fear of giving birth vaginally especially in the first pregnancy. The analysis suggested that fear of giving birth vaginally for some women was reinforced by difficult birth experience that was shown by the mother of pregnant woman.^25^

The folk tradition also reveals that the service providers usually prefer C-section because the profit margin in C-Section as compared to normal delivery is on higher side. Emotional attachment with first pregnancy usually creates a fragile atmosphere which allowed the health provider to offer more confidently for C-Section. This also unveils that hidden fear of unknown usually faced by the married couple. More importantly, this trend also score more when it comes to talking about the delivery cases among the educated couples as well as the cases being administered at wives’ paternal side. Fear from childbirth may vary among women. Some women would overcome and cope well with her fear, while others may have an extreme fear or anxiety; this may leads her to cope with her fear by request for caesarean section without medical indications.^26^Lack of second opinion consultation was found to affect physicians’ decision toward route of delivery. As it was found in the qualitative study conducted to determine factors affect obstetricians’ decision on the route of delivery among 19 boards certified obstetricians working in four Jordanian hospitals. The findings of the study showed that 100% of those obstetricians claimed that absence of second opinion consultation especially in the high risk deliveries plays a significant role in the increase in the C/S rate.^27^

### Study Limitations

- As women and family members were interviewed after the pregnancy outcomes, so a possibility of careful client responses cannot be ruled out.
- Also service providers at public health facilities were interviewed (many of these are known to be doing private practice in the evening), so there is possibility that views/ experiences of private providers are not fully reflected.

## Conclusion

Mother-in-law judgment on birthing matters is respected for some valid reasons, mainly related to their wider social experience and family trust. This suggests that any intervention for changing the safe birthing behaviour/ preference may include mother-in-law as a major stakeholder. The programme may include channels to address the socio-cultural issues linked with the consent of mother in law and religious importance of husbands.

Pregnant women consult their husbands and mother in laws for taking birthing decisions, the consultation with the skilled birth attendant for management of complication needs to be strengthened but requires more cultural freedom and mobility to it. Thus, the advice by skilled birth attendance affects in-time access to EmONC services.

Private clinics are preferred for various social and logistic reasons, however many private clinics doing caesarean are not adequately equipped. It is important to strengthen and engage private setups for EmONC as many fail to access public facility.

The Male reluctance (for cultural reasons) to make in-advance transport arrangement for possible EmONC is valid because to arrange a transport on a short notice is generally not a problem, however the travel voucher has limited utility if not covering the range of EmONC facilities. So, the idea of in-advance travel arrangements needs to be reviewed for cultural suitability and also to simplify the travel voucher process.

Unskilled birth attendants seem to have social preference as well as no evidence of their causing more neonatal and maternal deaths as compared to other attendants, so the question is whether to exclude or include with more support the traditional birth attendant for safer birthing. The exclusion of unskilled birth attendance needs to be reviewed in local context.

It can safely be concluded and recommended that any such health intervention requires inclusion of social sensitization and awareness component that effectively addresses the misconceptions among the targeted population. Similarly, there is an essential need of pre-project or pre-intervention strategy that advocates the inclusion of culturally sensitive staff of social scientists having ample knowledge of cultural patterns as well as taboos that may hinder the achievement of targets planned by such projects.

## Data Availability

All data produced in the present study are available upon reasonable request to the authors

